# The impact of timely cancer diagnosis on age disparities in colon cancer survival in New Zealand

**DOI:** 10.1101/2020.09.07.20189787

**Authors:** Sophie Pilleron, Camille Maringe, Hadrien Charvat, June Atkinson, Eva Morris, Diana Sarfati

**Affiliations:** Dept of Public Health, School of medicine, University of Otago, Wellington, New Zealand; Nuffield Department of Population Health, University of Oxford, Big Data Institute, Old Road Campus, Oxford, OX3 7LF, UK; Inequalities in Cancer Outcomes Network, London School of Hygiene and Tropical Medicine, Keppel Street, London WC1E 7HT, UK; Epidemiology and Prevention Group, Center for Public Health Sciences, National Cancer Center, Tokyo, Japan.; Section of cancer surveillance, International Agency for Research on Cancer, Lyon, France

**Keywords:** colon, cancer, survival, older adults, aged, population-based cancer registry, observational data

## Abstract

**Objective:** We described the role of patient-related and clinical factors on age disparities in colon cancer survival among patients aged 50-99 using New Zealand population-based cancer registry data linked to hospitalization data.

**Design:** We included new colon cancer cases diagnosed between 1 January 2006 and 31 July 2017, followed up to 31 December 2019. We linked these cases to hospitalisation data for the five years before the cancer diagnosis. We modelled the effect of age at diagnosis, sex, deprivation, comorbidity, and route to diagnosis on colon cancer survival by stage at diagnosis (localized, regional, distant, missing).

**Results:** Net survival decreased as the age at diagnosis increased, notably in advanced stages and missing stage. The excess mortality in older patients was minimal for localised cancers, maximal during the first six months for regional cancers, the first 18 months for distant cancers, and over the three years for missing stages. The age pattern of the excess mortality hazard varied according to sex for distant cancers, the route to diagnosis for regional and distant cancers, and comorbidity for cancer with missing stages.

**Conclusion:** The present population-based study shows that factors reflecting timeliness of cancer diagnosis most affected the difference in survival between middle-aged and older patients, probably by impacting treatment strategy. Because of the high risk of poor outcomes related to treatment in older patients, efforts made to improve earlier diagnosis in older patients are likely to help reduce age disparities in colon cancer survival in New Zealand.

## Introduction

Older patients with colon cancer have poorer cancer survival than middle-aged patients^1,2^. The disadvantage in cancer survival in older adults has even been widening^3,4^, suggesting that they have not benefitted from the improvements in cancer diagnosis and treatment at the same magnitude as younger people. Previous work showed that low survival in older patients with colon cancer was mainly explained by a high excess hazard in the first year after diagnosis^1^.

Cancer management in older patients may be complex. Because of age-related physiological changes, comorbidity, and polypharmacy, older patients have a higher risk of drug interaction, chemotherapy-related toxicity, and chemotherapy-induced peripheral neuropathy^5^. They are also more likely to have comorbidities^6^, cognitive impairments^7^, to be depressed^8^, and isolated with poor social support^9^. In addition, older patients are seldom included in randomized clinical trials^10^. Physicians have, therefore, to extrapolate from evidence based on younger patients to adapt new treatment strategies to older patients. As a consequence, older adults with cancer are likely to receive suboptimal treatment^11,12^. Besides, older patients are more likely to be diagnosed with cancer after an emergency presentation, and this has been associated with poor cancer survival^13^.

At the population level, older patients with colon cancer do not always have poorer survival than younger patients^14^. The magnitude of the age disparity in colon cancer relative survival varied inconsistently by stage at diagnosis, sex, ethnicity, and socioeconomic factors across studies^14^. While age is an important prognostic factor, only a few population-based studies described the role of patient and clinical factors on age disparities in colon cancer survival per se^14^.

In New Zealand, colon cancer was the second most common cancer and the second biggest cause of cancer deaths in adults aged 70 years or older in 2018^15^. The most recent 5-year net survival estimates for adults with colon cancer was 62% (95% confidence interval: 61%-63%)^16^, similar to most western countries^16,17^.

Using New Zealand population-based cancer registry data linked to hospitalization data, we describe, for the first time, the contribution of sex, ethnicity, deprivation, comorbidities, and route of diagnosis on age disparities in colon cancer survival.

## Methods

### Data sources

We included incident cases of colon cancer (ICD-10 codes C18.0-C18.9) diagnosed between 1 January 2006 and 31 July 2017 registered in the New Zealand population-based cancer registry (NZ-PBCR). We linked these cancer cases to hospitalisation data (National Minimum Dataset) for the five years before the cancer diagnosis, and to the outpatient dataset (National Non-Admitted Patient Collection) where all emergency admissions are recorded. We obtained the date of death for all cancer cases who died between 1 January 2006 and 31 December 2019 from the Ministry of Health.

### Inclusion criteria

We restricted our analyses to patients aged between 50 and 99 years old at diagnosis with a first occurrence of colon cancer. Colon cancer features and management among younger adults are different compared with cancer diagnosed at older ages^18,19^.

### Cancer-related data

We categorised the extent of disease as given by NZ-PBCR into Surveillance, Epidemiology and End Results (SEER) stage groups as follows: Localised to the organ of origin or Invasion of adjacent tissue or organ into SEER localised stage; Regional lymph nodes involvement into SEER regional stage; Distant into SEER distant stage; Not known into missing stage^20^.

### Clinical data

We used the “all-sites” weighted C3 index developed by Sarfati et al. to assess comorbidity among patients with cancer using administrative hospitalisation data^21^. We defined emergency presentation at diagnosis as a cancer diagnosis occurring in the 28 days following admission to an emergency department^22^.

### Socio-demographic data

Sex and age at diagnosis were available within the NZ-PBCR, but they originated from pathology reports and National Health Index. This information was available for all colon cancer cases. We used the New Zealand Deprivation Index (NZDep) to assess the socio-economic deprivation at cancer diagnosis for all cases^23^. The NZDep is an ecological indicator built using the nine following variables from the national census: housing tenure, benefit receipt, unemployment, income, telephone access, car access, single-parent families, education and household crowding. The NZDep was assigned to domicile code (census area) of each patient at the time of diagnosis. For all diagnoses occurring during the period 2006-2011, we used the NZDep based on information collected at the 2006 census; for all diagnoses occurring from 2012, we used the score based on information from the 2013 census. We could not map domicile code to an NZDep for 36 patients; we, therefore, excluded these cases from analyses.

Self-reported ethnicity available in the NZ-PBCR originated from the health records. We grouped patients into Māori and non-Māori. Ethnicity refers to the ethnic group people identify with or feel they belong to, and not to race, ancestry, nationality or citizenship (https://www.stats.govt.nz/topics/ethnicity). The ethnicity was not available for 249 patients; we, therefore, excluded these cases from analyses.

### Statistical analysis: Net survival estimation

When studying the survival of cancer patients, it is important to recognize they might die from their disease but also from other causes. This becomes crucial when looking at the effect of age at diagnosis as older patients are subjected to a greater risk of dying from competing causes (cardiovascular diseases, etc.). Net survival is a particularly interesting indicator in this context: it represents the hypothetical survival that would be observed if cancer were the only cause of death. Net survival is the indicator commonly used to analyse population-based cancer registry data when information on cause of death is either missing or deemed unreliable.

Net survival is derived from the estimation of individual excess mortality hazard (EMH): the overall hazard of death for cancer patients is assumed to be the sum of the expected (or background) hazard derived from population lifetables and the excess hazard due to cancer either directly (e.g., multiple organ failure resulting from cancer progression) or indirectly (e.g., treatment toxicity, pulmonary embolism). We used lifetables of mortality in the general population, defined by calendar period (2005-2007 and 2012-2014), single year of age, sex, and ethnicity (Māori, non-Māori) obtained from Statistic New Zealand. We built lifetables for each calendar year between 2006 and 2013 assuming linear interpolation between the age-, sex- and ethnic-specific mortality rates in 2005-7 and 2012-4. We extrapolated the rates beyond 2014 using the same constant yearly change in mortality. We fitted flexible excess hazard regression models for each stage at diagnosis. Since we are interested in patterns of mortality in the first three years after diagnosis, we censored all patients who were still alive beyond five years after diagnosis for model stability at the tails^24^.

### Model-building strategy

We selected the best-fitting model for each stage at diagnosis. First, we selected the functional form of the baseline hazard on the most complex model yielding minimal Aikake Information Criterion (AIC). For regional, distant and missing stages, the functional form was chosen among the exponential of a restricted cubic spline and the exponential of B-spline of degree 2 or 3; for each option, we tested the number of knots (one or two) located at the median or at the tertile of the distribution of survival times in patients who died. For patients with localised stage at diagnosis, the model did not converge using any of these forms, so we eventually selected the best fitting functional form among piecewise constant hazards with 0, 1 or 2 knots located as previously described. Once the functional form of the baseline hazard defined, we selected our final models using the model building strategy of Wynant and Abrahamowicz^25^, adapted for relative survival setting by Maringe et al.^26^. We forced the lifetable variables (i.e. sex, age, ethnicity, year at diagnosis) into the models as recommended^27–29^. We tested the non-linear effect of age at diagnosis, year of diagnosis and comorbidity score by constructing and modelling the effects of restricted cubic splines with a knot located at the median of their distributions and boundary knots at the 10^th^ and 90^th^ percentiles of their distributions. For all covariates, we tested for time-dependent effects and their interaction with age at diagnosis using the Likelihood Ratio Test.

All predictions used in all Figures in the manuscript were made for Non-Māori males diagnosed in 2012, who lived in an area with third quintile of deprivation index, with median comorbidity score, no emergency admission recorded in the month prior to their cancer diagnosis.

We performed data management using Stata (version 16.0; StataCorp, 2019) and statistical analyses using R statistical software (version 3.4.0; R Development Core Team, 2017), in particular the ‘mexhaz’ package was used for flexible excess hazard modelling^30^.

## Results

Out of 23,003 patients diagnosed with colon cancer between January 2006 and July 2017, we included 21,270 patients aged 50-99 (median age at diagnosis = 74, interquartile range 67-81; 51.7% were females). Table 1 presents the characteristics of the included patients by age at diagnosis, and supplementary Table 1 by age at diagnosis and stage at diagnosis. While patients less than 75 years old were mostly males, older patients were mainly females. Non-Māori constituted the majority of the newly diagnosed patients, and their percentage increased as the age at diagnosis increased. Deprivation level was almost evenly distributed across age groups. Overall, 24.0% of patients were diagnosed with localised cancer, 43.2% with regional cancer, 21.3% with distant cancer, and 11.4% had a missing stage. The percentage of missing stage increased after the age of 75, and it was twice higher in the 85-99 age group than in the 75-84 age group. The comorbidity score increased with chronological age, with over three quarters of patients aged 85 years or over who had a positive comorbidity score compared to half of patients aged 50-64. Overall, 29.7% of patients were diagnosed with colon cancer after an admission at emergency department within the previous 28 days, and the percentage was the highest in the oldest age group.

**Table 1.** Patients’ characteristics by age group

| Age categories (years) | 50-64 | 65-74 | 75-84 | 85-99 |
| --- | --- | --- | --- | --- |
| Cases (%) | 4212 (19.8) | 6614 (31.1) | 7371 (34.7) | 3073 (14.4) |
| Male (%) | 2193 (52.1) | 3499 (52.9) | 3458 (46.9) | 1126 (36.6) |
| Ethnicity (%) |  |  |  |  |
| Māori | 384 (9.1) | 335 (5.1) | 208 (2.8) | 43 (1.4) |
| Non- Māori | 3828 (90.9) | 6279 (94.9) | 7163 (97.2) | 3030 (98.6) |
| Deprivation Index quintiles (%) |  |  |  |  |
| 1 - Less deprived | 831 (19.7) | 1182 (17.9) | 1133 (15.4) | 463 (15.1) |
| 2 | 782 (18.6) | 1244 (18.8) | 1353 (18.4) | 553 (18.0) |
| 3 | 873 (20.7) | 1450 (21.9) | 1673 (22.7) | 700 (22.8) |
| 4 | 946 (22.5) | 1587 (24.0) | 1882 (25.5) | 816 (26.6) |
| 5 - Most deprived | 780 (18.5) | 1151 (17.4) | 1330 (18.0) | 541 (17.6) |
| Stage at diagnosis (%) |  |  |  |  |
| Localised | 1020 (24.2) | 1712 (25.9) | 1854 (25.2) | 529 (17.2) |
| Regional | 1782 (42.3) | 3060 (46.3) | 3261 (44.2) | 1087 (35.4) |
| Distant | 1115 (26.5) | 1392 (21.0) | 1405 (19.1) | 619 (20.1) |
| Missing | 295 ( 7.0) | 450 ( 6.8) | 851 (11.5) | 838 (27.3) |
| Comorbidity score >0 (%) | 2159 (51.3) | 4123 (62.3) | 5394 (73.2) | 2408 (78.4) |
| Comorbidity score (median [IQR]) | 0.39 [0.00,1.27] | 0.75 [0.00,1.89] | 1.21 [0.00,2.71] | 1.38 [0.52,2.81] |
| Emergency route (%) | 1225 (29.1) | 1746 (26.4) | 2164 (29.4) | 1176 (38.3) |

IQR: Inter-quartile range

The estimated 1-, and 3- year net survival were, respectively, 98.1% (95% confidence interval: 97.6%-98.5%), and 95.8% (95.0%-96.6%) in patients with localised cancer, 91.4% (90.7%-92.0%), and 79.8% (78.8%-80.8%) in patients with regional cancer, 38.3% (37.0%-39.6%), and 15.9% (14.9%-17.0%) in patients with distant cancer, and 55.2% (53.3%-57.1%), and 35.1% (33.1%-37.0%) in patients with missing stage. Net survival decreased as the age a diagnosis increased in all stages, but the disadvantage in older patients was more at later stages of cancer diagnosis and for cancer with missing stage (Figure 1, supplementary Table 2).

**Figure 1.**
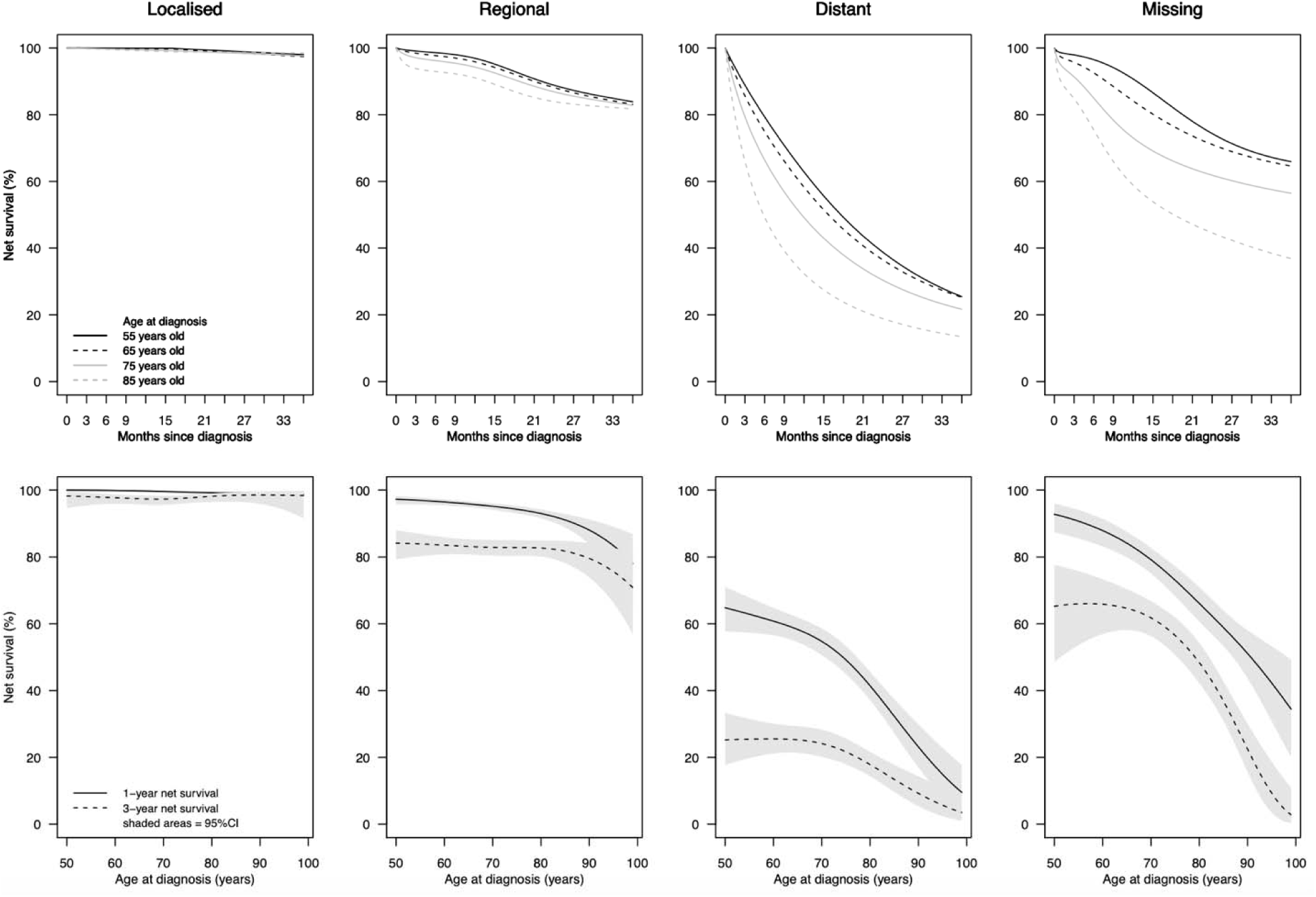
Net survival for patients aged 55, 65, 75 and 85 years old over the first three years after diagnosis by stage at diagnosis (top panel) and one-and three-year net survival by age at diagnosis and stage at diagnosis (bottom panel)

Figure 2 shows one-year net survival by age at diagnosis, stage at diagnosis and patient-related and clinical factors. One-year net survival did not differ much by sex for patients with localised cancers. For regional stage, one-year net survival was similar across sexes up to 80 years of age, then female patients had better net survival than males. For distant stages, net survival in female patients decreased almost linearly as age increased, while it is not the case for male patients. Male patients aged 65-85 years old had better survival than females. Females with missing stage had lower net survival than males. Māori patients with localised cancer had similar net survival than non-Māori patients regardless of the age at diagnosis. For the other stages, Māori patients had poorer net survival than Non-Māori patients. For cancers with regional and distant spread, net survival decreased as the deprivation index increased, but the difference across deprivation index quintiles was small, and similar over time. Net survival decreased as comorbidity score increased in patients with localised and regional cancers. However, patients with higher level of comorbidity had better survival than those with lower level in patients with distant cancer. For patients with missing stage, patients with high comorbidity level had poorer survival than other patients up to the age of 70 years old. Afterwards, they had better one-year net survival. Patients diagnosed with colon cancer after an emergency admission had poorer net survival regardless of the stage or the age at diagnosis. The difference in one-year net survival between those who had an emergency presentation and those who had not increased as age increased, except for patients with distant cancer for whom the reverse was observed. Similar results are observed three years after diagnosis (Supplementary Figure 1), with a few exceptions. Māori patients with distant cancer had slightly better three-year net survival than non-Māori patients. Difference in net survival across comorbidity levels were marked in patients with localised and regional cancers at three than at one year since diagnosis. Patients with higher comorbidity level had poorer three-year net survival than others when cancer was diagnosed at later stage. For patients with missing stage, patients with higher comorbidity level had significantly poorer net survival than other but differences disappeared

**Figure 2.**
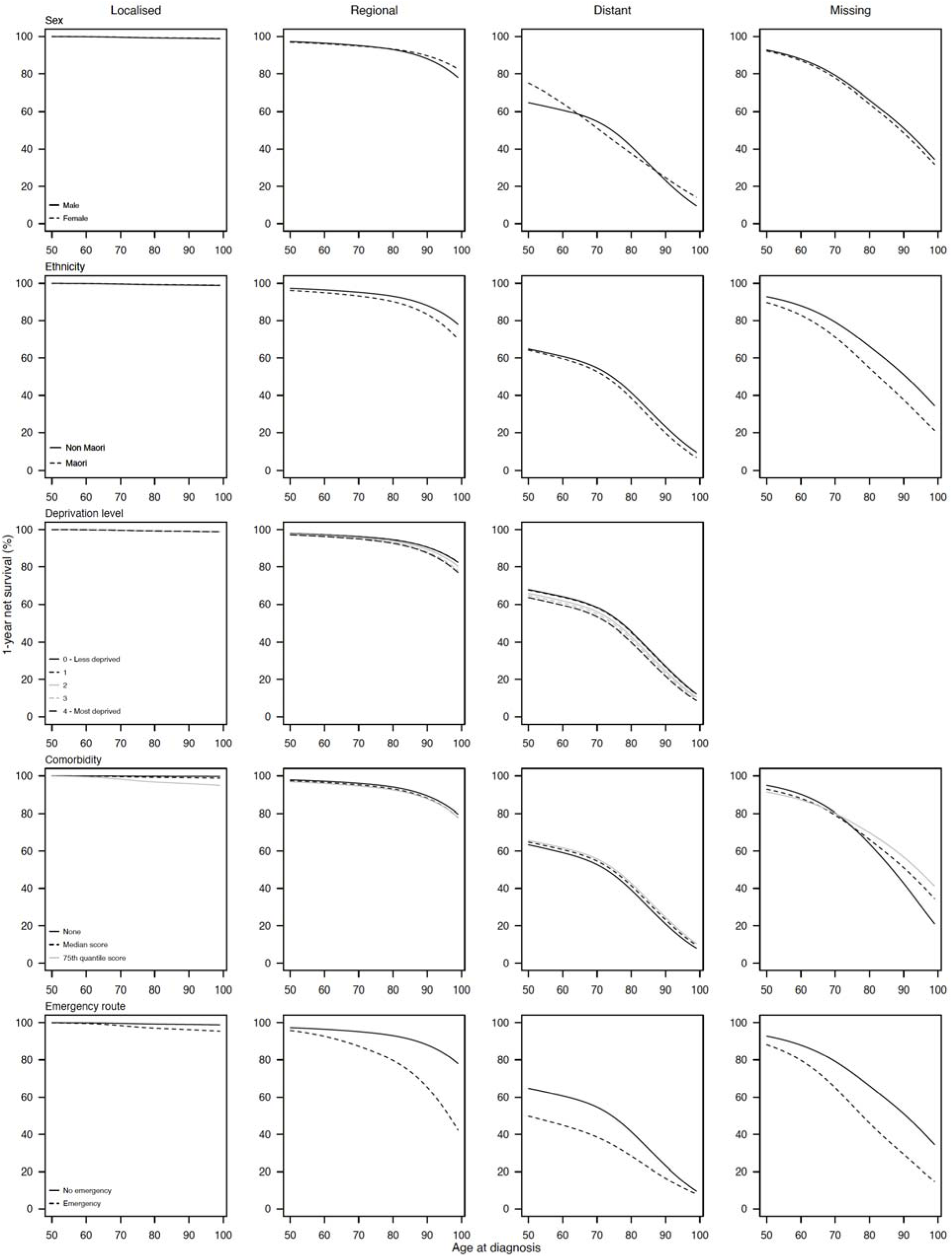
One-year net survival by age at diagnosis, stage at diagnosis and patient-related and clinical factor

To better understand the age difference in net survival, we looked at the pattern of excess mortality hazard over time by age at diagnosis (Figure 3). For patients with localised cancer, the EMH was low for all ages, with a sligh disadvantage for older patients during the first 15 months after diagnosis. The tendency was reversed afterwards. Fo regional cancers, the excess mortality in older patients was obvious during the first six months after diagnosis. Then the EMHs were quite similar across all ages, the EMH ratios being close to 1. For distant cancers, older patients had higher excess mortality than younger patients for 18 months after diagnosis. Then, the curves converged for all ages and the EMH ratios were close to 1. For patients with missing stage, the excess in mortality in older adults was highest during the first year after diagnosis and persisted over the three years of follow-up for the oldest patients.

**Figure 3.**
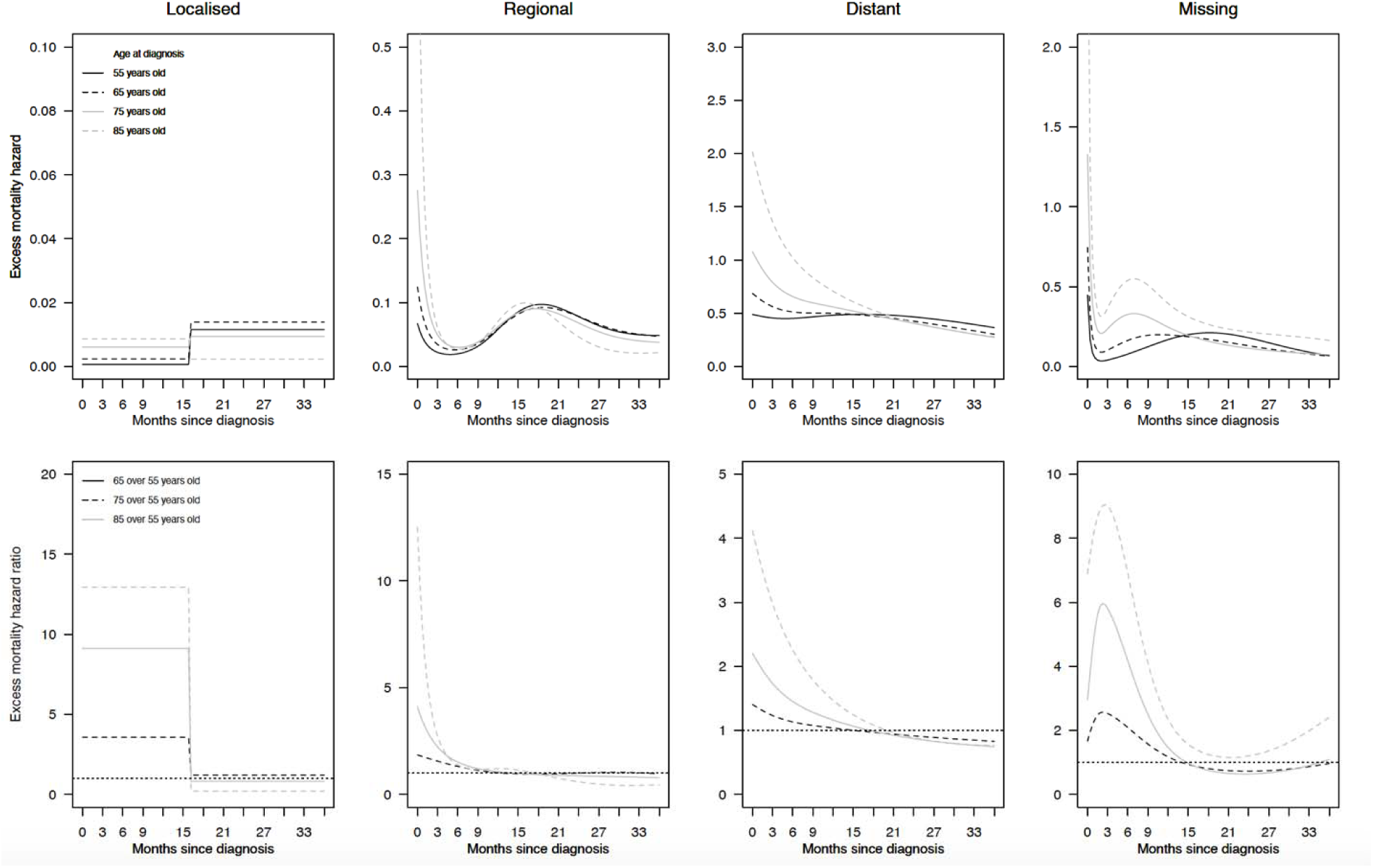
Excess mortality hazard for patients aged 55, 65, 75 and 85 years old (top panel), and excess mortality hazard ratio for patients aged 65, 75 and 85 years compared to patients aged 55 years old during the first three years since diagnosis by stage at diagnosis (bottom panel)

We tested the modification effect of each factor on the relationship of age at diagnosis on the EMH (supplementary Table 3). For patients with localised cancer, no factors modified the effect of age at diagnosis on the EMH. For cancers with regional spread, the effect of age at diagnosis on the EMH was different based on the route to diagnosis, with greater EMH ratio in patients diagnosed after an emergency admission than other patients, especially in the first three months after diagnosis (Figure 4-A). For cancer with distant spread, the pattern of EMH by age at diagnosis varied across sexes and route to diagnosis (Figure 4 – B and C). Patients who were not diagnosed after an emergency presentation had higher EMH ratios than those who were diagnosed through emergency settings. This difference was visible mainly in the first six months (Figure 4 –B). The EMH ratio reached 1 after the first year for patients with an emergency presentation, and later for the other patients. Females had higher excess mortality hazard ratios compared to males over the entire follow-up time. However, the sex difference was greatest in the first three months after diagnosis and decreased afterwards to become negligible at three years (Figure 4- C). For cancer with missing stage, comorbidity level modified considerably the effect of age at diagnosis on the EMH, with EMH ratios of up to 10 for patients with 25^th^ percentile comorbidity score. The lower comorbidity score, the higher the EMH ratio, mainly in the first six months after diagnosis and in patients older than 80 years old (Figure 4-D).

**Figure 4.**
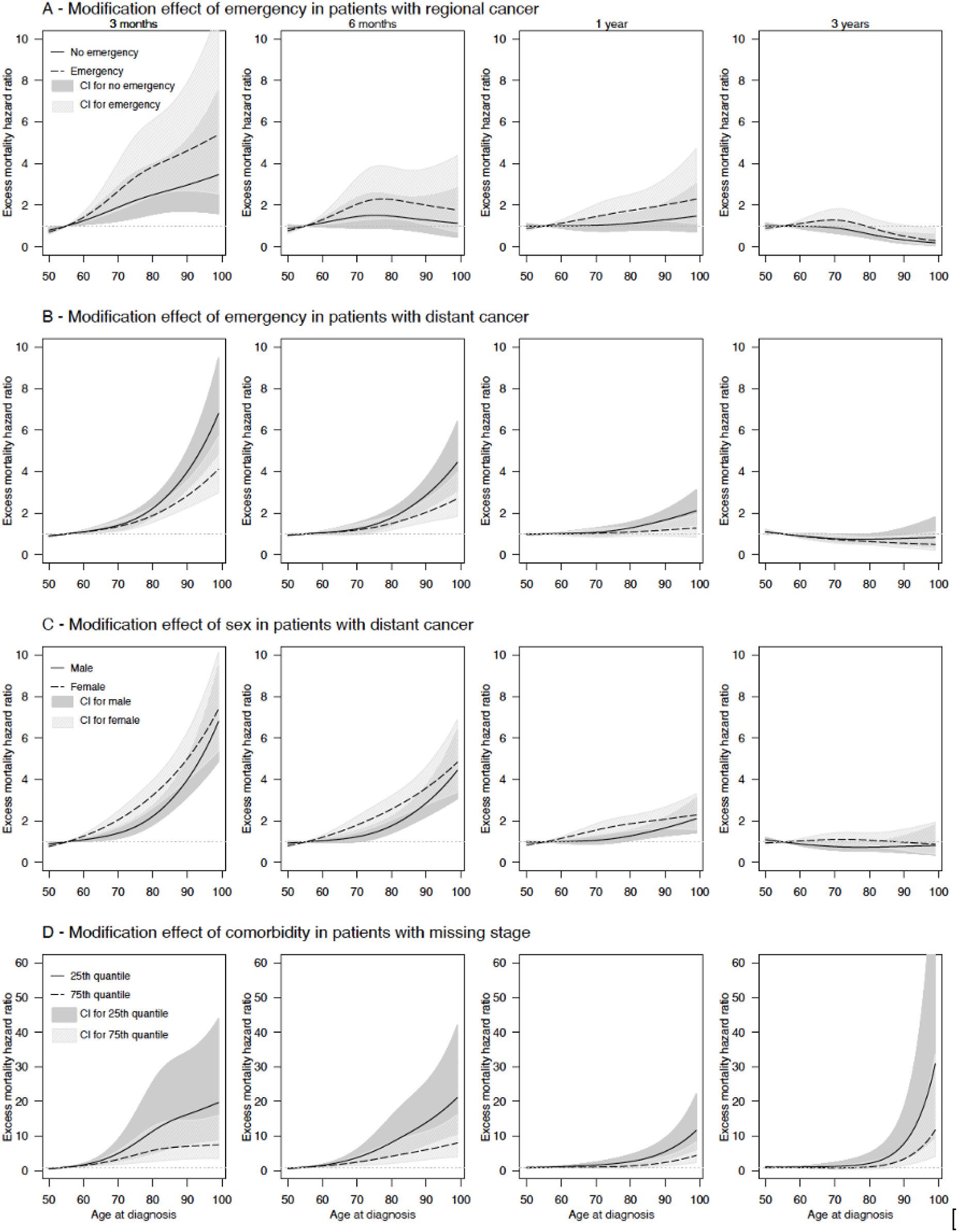
Excess mortality hazard ratios at 3, 6 months,1 and 3 years by age at diagnosis based on (A) route to diagnosis in patients with regional colon cancer and (B) those with distant colon cancer, (C) sex in patients with distant colon cancer and (D) comorbidity in patients with missing stage (reference: 55 years old)

## Discussion

This study, for the first time, provides a comprehensive description of patient-related and clinical factors that influence the effect of age at diagnosis on the excess mortality hazard in patients with colon cancer aged 50 years or older in New Zealand. We found that the stage at diagnosis and an emergency presentation at diagnosis modified the most the age disparity in colon cancer survival, while comorbidity played little role. About patient-related factors, sex was the sole factor that influenced age disparities in cancer survival but only in patients with distant cancer, females having greater age difference in net survival than males.

The female advantage in colon cancer survival in patients aged below 65 years old observed in our study is congruent with findings from the EUROCARE study^31^. In Finland, the absolute difference in one-year relative survival between the 45-59-year group and the 75-and-over group was also higher in females compared to males with distant colon cancer^32^. As suggested by other authors, the interaction between the age at diagnosis and sex indicates possible involvement of sex hormones^31,33^. However, the causal mechanism explaining this interaction and the reason why this concerns only patients with distant cancer remain unclear and deserve further investigation.

We are not aware of other studies looking at the effect of ethnicity on age disparities in colon cancer survival. However, one U.S. study showed black patients with colorectal cancer had greater age disparity in five-year net survival than white patients, mainly explained by poorer survival among black patients aged 75 or over compared to white patients^34^. Because the relationship between ethnicity and healthcare system is very complex and specific to each country’s history, any comparisons across countries are not relevant. While our study confirmed the negative effect of deprivation on colon cancer survival^35^, we did not find any evidence of its role in age disparities in colon cancer survival.

Comorbidity is highly prevalent in patients with colon cancer, especially in older patients^36^, and may affect the likelihood to receive treatment^37^, and impact survival^38^. However, our results suggest that comorbidity does not fully explain the disparity in survival observed between older and middle-aged patients with colon cancer in New Zealand. In other terms, for cancers with known stage, the effect of age on the excess hazard of death was the same across all comorbidity levels. In our study, comorbidity was measured using hospitalisation data that may not capture all aspects of comorbidity, especially in patients with comorbidity that did not require hospitalisation. However, in New Zealand, the C3 index performed similarly or even slightly better than a comorbidity index based on pharmaceutical data, that identify patients who had drug prescription for chronic diseases irrespective of whether the patient was hospitalised or not^39^. Our results may also suggest that the presence of comorbidity affects cancer management, regardless of the age at diagnosis. However, a difference in treatment based on the chronological age or the existence of other unmeasured factors may explain lower survival in older patients without comorbidity. Indeed, comorbidity alone does not reflect the overall fitness status of older patients. As older patients are heterogeneous in terms of health status and fitness, the comprehensive assessment of common geriatric conditions including functional status, falls, cognition, nutritional status, social support helps identifying patients who may benefit the most from cancer treatment^40^. However, to date, such information is not currently available in the cancer registry or hospitalisation datasets.

Factors related to timeliness of cancer diagnosis — i.e. the stage at diagnosis and an emergency presentation — greatly affected age disparities in colon cancer survival. This observation reflects the effect of cancer management on survival. The treatment phase is a critical period where age inequalities occur^41^, likely because physicians are lacking evidence-based treatment strategies, especially in comorbid and oldest patients^42^. Even though surgery is the standard treatment in early-stage colon cancers^43,44^, older patients have a reduced likelihood of undergoing surgery^45–47^. They also experienced higher post-operative mortality rates up to one year after surgery^48^. In New Zealand, a recent report on colon cancers (all stages combined) diagnosed during the 2013-2016 period showed that 64% of patients aged 75 years or older had a major surgery that was similar to 66% in the 60-74 age group, and higher than in younger patients (58%-59%). The percentage of death within 90 days of resection was 7.3% in patients aged 75 or over, and 4.8% after elective resection against 1.5-2.9% and 0.0-1.7%, respectively, in patients aged under 75^49^. Older patients with colon cancer are also less likely to receive chemotherapy^50–52^. In New Zealand, patients with colon cancer aged 75 or over were nine-times less likely to receive chemotherapy than those aged 25–54 years old^37^. Yet, evidence suggests that fit older patients may benefit from the same regimen of chemotherapy as middle-aged patients^53,54^, and less intensive therapies may be used in unfit older patients^55^. Consistently with previous studies^13^, we showed that older adults were more prone to be diagnosed through emergency settings than middle-aged patients. An emergency presentation at diagnosis was associated with a lower chance of curative treatment and an excess risk of mortality, particularly in the initial months after diagnosis^13^. The current New Zealand guidelines for non-metastatic colon cancer management do not include recommendations regarding chemotherapy or surgery tailored to older patients^43^. Yet, without appropriate stratification, older patients are more likely to receive suboptimal treatment compared to middle-aged patients. Further observational studies to investigate how treatment may influence age disparities in colon cancer survival at population-level are needed. However, these studies should be properly designed to remove the risk of immortal-time bias, arising when the time of treatment initiation does not coincide with time of cancer diagnosis^56^.

Because of the high risk of poor outcomes related to treatment in older patients, effort should be made on reducing delays in colon cancer diagnosis. The roll-out of the national colorectal screening program began in 2017. However, the screening program will likely increase age disparities in survival because it is targeting only adults aged 60-74 years old; older patients will not benefit from it. Besides, unfortunately, many colon cancer symptoms, including abdominal pain, diarrhoea, and constipation, have low specificity^57^ and are prevalent in older patients, especially in individuals with other comorbid conditions. However, safety-netting approach –i.e. planned re-evaluation of vague symptoms - may help to detect cancer earlier^58^. Evidence suggests that patient-related, social, clinical, or health-system-related factors may cause delays in cancer diagnosis in older patients^59,60^. At the patient level, presence of comorbidity increased the likelihood to be diagnosed with distant colorectal cancer in New Zealand^61^. However, tumour stage was not correlated with the delay from the onset of symptoms to the administration of definite treatment^62^. Still, the potential effect of age was not analysed, and the audit was restricted to one hospital only^62^. It is also possible that older people are more likely to face barriers to timely accessing health care because of socioeconomic factors^63^. A comprehensive understanding of factors influencing timely diagnosis of colon cancer in older adults is warranted in New Zealand to develop interventions to improve earlier cancer diagnosis, regardless of age, that will ultimately reduce age disparities observed at the population level.

Our study has limitations. The estimation of net survival relies on the background mortality rates obtained from lifetables. The use of net survival implies that one matches patients with cancer to individuals from the general population that should ideally present the same characteristics (sex, age, socio-economic level, comorbidity, etc.) as patients except for cancer. When one is interested in the effect of age on cancer survival, some other characteristics, such as comorbidity, that influences both background mortality and cancer mortality, have their importance. However, this information was unavailable at population level. Consequently, the estimation of excess mortality hazards may be overestimated, and therefore our net survival estimates underestimated^64^.

Finally, patient-related and clinical factors may have different effects on age disparities in cancer survival in other high-income countries because of differences in the health-care system. Similar studies in other countries would be interesting to test the generalizability of our findings.

## Conclusion

The present population-based study shows that factors reflecting timeliness of cancer diagnosis affected the most the difference in colon cancer survival between middle-aged and older patients, probably due to their impact on treatment strategy. In contrast, comorbidity and patient-related factors play a negligible role in age disparities in colon cancer survival in New Zealand. It is Utopian to believe that colon cancer survival in older patients may equal that in middle-aged patients. However, our results suggest there are opportunities for enhancement, notably in improving earlier diagnosis in older adults. Efforts towards this goal are likely to help reduce age disparities in colon cancer survival in New Zealand.

## Data Availability

The data that support the findings of this study are available from New Zealand Ministry of Health. Restrictions may apply to the availability of these data outside New Zealand. See https://www.health.govt.nz/nz-health-statistics/access-and-use/data-request-form.

## Acknowledgment

We thank New Zealand cancer registry staff for their fantastic work and their help in understanding cancer data. We also thank Kendra Telfer for her immeasurable help in understanding NZ datasets and Dr Nicholas West for answering our questions about cancer pathology.

## Disclaimer

Where authors are identified as personnel of the International Agency for Research on Cancer/World Health Organization, the authors alone are responsible for the views expressed in this article and they do not necessarily represent the decisions, policy or views of the International Agency for Research on Cancer/World Health Organization.

## References

1. Colonna, M., Bossard, N., Remontet, L., Grosclaude, P. & FRANCIM Network. Changes in the risk of death from cancer up to five years after diagnosis in elderly patients: a study of five common cancers. Int. J. Cancer 127, 924–931 (2010).

2. Quaglia, A. et al. The cancer survival gap between elderly and middle-aged patients in Europe is widening. Eur. J. Cancer Oxf. Engl. 1990 45, 1006–1016 (2009).

3. De Angelis, R. et al. Cancer survival in Europe 1999-2007 by country and age: results of EUROCARE--5-a population-based study. Lancet Oncol. 15, 23–34 (2014).

4. Arnold, M. et al. Progress in cancer survival, mortality, and incidence in seven high-income countries 1995–2014 (ICBP SURVMARK-2): a population-based study. Lancet Oncol. 0, (2019).

5. van Abbema, D. L. et al. Patient- and tumor-related predictors of chemotherapy intolerance in older patients with cancer: A systematic review. J. Geriatr. Oncol. 10, 31–41 (2019).

6. Yancik R. et al. Comorbidity and age as predictors of risk for early mortality of male and female colon carcinoma patients: A population-based study. Cancer 82, 2123–2134 (1998).

7. Vega, J. N., Dumas, J. & Newhouse, P. A. Cognitive Effects of Chemotherapy and Cancer-Related Treatments in Older Adults. Am. J. Geriatr. Psychiatry 25, 1415–1426 (2017).

8. Weinberger, M. I., Bruce, M. L., Roth, A. J., Breitbart, W. & Nelson, C. J. Depression and barriers to mental health care in older cancer patients. Int. J. Geriatr. Psychiatry 26, 21–26 (2011).

9. Williams, G. R. et al. Unmet social support needs among older adults with cancer. Cancer 125, 473–481 (2019).

10. Hutchins, L. F., Unger, J. M., Crowley, J. J., Coltman, C. A. & Albain, K. S. Underrepresentation of patients 65 years of age or older in cancer-treatment trials. N. Engl. J. Med. 341, 2061–2067 (1999).

11. Foster, J. A., Salinas, G. D., Mansell, D., Williamson, J. C. & Casebeer, L. L. How does older age influence oncologists’ cancer management? The Oncologist 15, 584–592 (2010).

12. Walter, V. et al. Decreasing Use of Chemotherapy in Older Patients With Stage III Colon Cancer Irrespective of Comorbidities. J. Natl. Compr. Cancer Netw. JNCCN 17, 1089–1099 (2019).

13. Zhou, Y. et al. Diagnosis of cancer as an emergency: a critical review of current evidence. Nat. Rev. Clin. Oncol. 14, 45–56 (2017).

14. Pilleron, S. et al. Patterns of age disparities in colon and lung cancer survival: a systematic narrative literature review. BMJ Open Submitt. (2020).

15. Ferlay, J. et al. Global Cancer Observatory. https://gco.iarc.fr/.

16. Araghi, M. et al. Colon and rectal cancer survival in seven high-income countries 2010–2014: variation by age and stage at diagnosis (the ICBP SURVMARK-2 project). Gut (2020) doi: 10.1136/gutjnl-2020-320625.

17. Allemani, C. et al. Global surveillance of trends in cancer survival 2000-14 (CONCORD-3): analysis of individual records for 37 513 025 patients diagnosed with one of 18 cancers from 322 population-based registries in 71 countries. Lancet Lond. Engl. 391, 1023–1075 (2018).

18. Willauer, A. N. et al. Clinical and molecular characterization of early-onset colorectal cancer. Cancer (2019) doi: 10.1002/cncr.31994.

19. Mauri, G. et al. Early-onset colorectal cancer in young individuals. Mol. Oncol. 13, 109–131 (2019).

20. SEER Training Modules, Staging a Cancer Case.U. S. National Institutes of Health, National Cancer Institute. (2020).

21. Sarfati, D. et al. Cancer-specific administrative data-based comorbidity indices provided valid alternative to Charlson and National Cancer Institute Indices. J. Clin. Epidemiol. 67, 586–595 (2014).

22. Elliss-Brookes, L. et al. Routes to diagnosis for cancer – determining the patient journey using multiple routine data sets. Br. J. Cancer 107, 1220–1226 (2012).

23. Salmond, C. E. & Crampton, P. Development of New Zealand’s deprivation index (NZDep) and its uptake as a national policy tool. Can. J. Public Health Rev. Can. Sante Publique 103, S7–11 (2012).

24. Remontet, L., Bossard, N., Belot, A., Estève, J. & French network of cancer registries FRANCIM. An overall strategy based on regression models to estimate relative survival and model the effects of prognostic factors in cancer survival studies. Stat. Med. 26, 2214–2228 (2007).

25. Wynant, W. & Abrahamowicz, M. Impact of the model-building strategy on inference about nonlinear and time-dependent covariate effects in survival analysis. Stat. Med. 33, 3318–3337 (2014).

26. Maringe, C., Belot, A., Rubio, F. J. & Rachet, B. Comparison of model-building strategies for excess hazard regression models in the context of cancer epidemiology. BMC Med. Res. Methodol. 19, 210 (2019).

27. Estève, J., Benhamou, E., Croasdale, M. & Raymond, L. Relative survival and the estimation of net survival: elements for further discussion. Stat. Med. 9, 529–538 (1990).

28. Perme, M. P., Stare, J. & Estève, J. On Estimation in Relative Survival. Biometrics 68, 113–120 (2012).

29. Danieli, C., Remontet, L., Bossard, N., Roche, L. & Belot, A. Estimating net survival: the importance of allowing for informative censoring. Stat. Med. 31, 775–786 (2012).

30. Charvat, H. & Belot, A. mexhaz: Mixed Effect Excess Hazard Models. R package version 1.5.

31. de Vries E. et al. The advantage of women in cancer survival: An analysis of EUROCARE-4 data. Eur. J. Cancer 45, 1017–1027 (2009).

32. Dickman, P. W. et al. Survival of cancer patients in Finland 1955-1994. Acta Oncol. Stockh. Swed. 38 Suppl 12, 1–103 (1999).

33. Quirt, J. S., Nanji, S., Wei, X., Flemming, J. A. & Booth, C. M. Is there a sex effect in colon cancer? Disease characteristics, management, and outcomes in routine clinical practice. Curr. Oncol. 24, e15–e23 (2017).

34. Wingo, P. A., Ries, L. A. G., Parker, S. L. & Heath, C. W. Long-term cancer patient survival in the United States. Cancer Epidemiol. Biomarkers Prev. 7, 271–282 (1998).

35. Jeffreys, M. et al. Socioeconomic Inequalities in Cancer Survival in New Zealand: The Role of Extent of Disease at Diagnosis. Cancer Epidemiol. Biomarkers Prev. 18, 915–921 (2009).

36. Edwards, B. K. et al. Annual report to the nation on the status of cancer, 1975-2006, featuring colorectal cancer trends and impact of interventions (risk factors, screening, and treatment) to reduce future rates. Cancer 116, 544–573 (2010).

37. Sarfati, D. et al. The effect of comorbidity on the use of adjuvant chemotherapy and survival from colon cancer: a retrospective cohort study. BMC Cancer 9, 116 (2009).

38. Sarfati, D., Koczwara, B. & Jackson, C. The impact of comorbidity on cancer and its treatment. CA. Cancer J. Clin. 66, 337–350 (2016).

39. Sarfati, D., Gurney, J., Stanley, J., Lim, B. T. & McSherry, C. Development of a Pharmacy-based Comorbidity Index for Patients With Cancer. Med. Care 52, 586–593 (2014).

40. Mohile, S. G. et al. Practical Assessment and Management of Vulnerabilities in Older Patients Receiving Chemotherapy: ASCO Guideline for Geriatric Oncology. J. Clin. Oncol. 36, 2326–2347 (2018).

41. Hayes, L. et al. Age-related inequalities in colon cancer treatment persist over time: a population-based analysis. J. Epidemiol. Community Health 73, 34–41 (2019).

42. Townsley, C. A., Selby, R. & Siu, L. L. Systematic review of barriers to the recruitment of older patients with cancer onto clinical trials. J. Clin. Oncol. Off. J. Am. Soc. Clin. Oncol. 23, 3112–3124 (2005).

43. New Zealand Guidelines Group. Clinical practice guidelines for the management of early colorectal cancer. (2011).

44. Papamichael, D. et al. Treatment of colorectal cancer in older patients: International Society of Geriatric Oncology (SIOG) consensus recommendations 2013. Ann. Oncol. Off. J. Eur. Soc. Med. Oncol. 26, 463–476 (2015).

45. Gooiker, G. A. et al. Risk Factors for Excess Mortality in the First Year After Curative Surgery for Colorectal Cancer. Ann. Surg. Oncol. 19, 2428–2434 (2012).

46. Dekker, J. W. T. et al. Cause of death the first year after curative colorectal cancer surgery; a prolonged impact of the surgery in elderly colorectal cancer patients. Eur. J. Surg. Oncol. J. Eur. Soc. Surg. Oncol. Br. Assoc. Surg. Oncol. 40, 1481–1487 (2014).

47. Benitez Majano, S. et al. Surgical treatment and survival from colorectal cancer in Denmark, England, Norway, and Sweden: a population-based study. Lancet Oncol. 20, 74–87 (2019).

48. Dekker J.W.T. et al. Importance of the first postoperative year in the prognosis of elderly colorectal cancer patients. Ann. Surg. Oncol. 18, 1533–1539 (2011).

49. Ministry of Health. Bowel cancer quality improvement report. (2019).

50. Merchant, S. J. et al. Management of stage III colon cancer in the elderly: Practice patterns and outcomes in the general population. Cancer 123, 2840–2849 (2017).

51. Raycraft, T. et al. Causes of mortality in older patients with stage 3 colon cancer. J. Geriatr. Oncol. 10, 138–142 (2019).

52. Sarasqueta, C. et al. Impact of age on the use of adjuvant treatments in patients undergoing surgery for colorectal cancer: patients with stage III colon or stage II/III rectal cancer. BMC Cancer 19, 735 (2019).

53. Taieb, J. How best to treat older patients with metastatic colorectal cancer? Lancet Gastroenterol. Hepatol. 4, 331–333 (2019).

54. Van Cutsem, E. et al. ESMO consensus guidelines for the management of patients with metastatic colorectal cancer. Ann. Oncol. Off. J. Eur. Soc. Med. Oncol. 27, 1386–1422 (2016).

55. Winther, S. B. et al. Reduced-dose combination chemotherapy (S-1 plus oxaliplatin) versus full-dose monotherapy (S-1) in older vulnerable patients with metastatic colorectal cancer (NORDIC9): a randomised, open-label phase 2 trial. Lancet Gastroenterol. Hepatol. 4, 376–388 (2019).

56. Maringe, C. et al. Reflections on modern methods: trial emulation in the presence of immortal-time bias. Assessing the benefit of major surgery for elderly lung cancer patients using observational data. Int. J. Epidemiol. (2020) doi: 10.1093/ije/dyaa057.

57. Koo, M. M., Hamilton, W., Walter, F. M., Rubin, G. P. & Lyratzopoulos, G. Symptom Signatures and Diagnostic Timeliness in Cancer Patients: A Review of Current Evidence. Neoplasia 20, 165–174 (2018).

58. Nicholson, B. D., Mant, D. & Bankhead, C. Can safety-netting improve cancer detection in patients with vague symptoms? BMJ 355, i5515 (2016).

59. Brousselle, A. et al. Explaining time elapsed prior to cancer diagnosis: patients’ perspectives. BMC Health Serv. Res. 17, 448 (2017).

60. Sikdar, K. C., Dickinson, J. & Winget, M. Factors associated with mode of colorectal cancer detection and time to diagnosis: a population level study. BMC Health Serv. Res. 17, 7 (2017).

61. Gurney, J., Sarfati, D. & Stanley, J. The impact of patient comorbidity on cancer stage at diagnosis. Br. J. Cancer 113, 1375–1380 (2015).

62. Tiong, J., Gray, A., Jackson, C., Thompson-Fawcett, M. & Schultz, M. Audit of the association between length of time spent on diagnostic work-up and tumour stage in patients with symptomatic colon cancer. ANZ J. Surg. 87, 138–142 (2017).

63. Vercelli, M. et al. Cancer survival in the elderly: effects of socio-economic factors and health care system features (ELDCARE project). Eur. J. Cancer Oxf. Engl. 1990 42, 234–242 (2006).

64. Rubio, F. J., Rachet, B., Giorgi, R., Maringe, C. & Belot, A. On models for the estimation of the excess mortality hazard in case of insufficiently stratified life tables. Biostatistics (2019) doi: 10.1093/biostatistics/kxz017.

